# Vitamin D deficiency increases vulnerability to canagliflozin-induced adverse effects on 1,25-dihydroxyvitamin D and PTH

**DOI:** 10.1101/2023.05.11.23289854

**Authors:** Zhinous Shahidzadeh Yazdi, Elizabeth A. Streeten, Hilary B. Whitlatch, May E. Montasser, Amber L. Beitelshees, Simeon I. Taylor

## Abstract

*Context.* Canagliflozin has been reported to increase the risk of bone fracture – possibly mediated by decreasing 1,25-dihydroxyvitamin D [1,25(OH)_2_D] and increasing PTH.

*Objective.* To investigate whether baseline vitamin D (VitD) deficiency renders individuals vulnerable to this adverse effect and whether VitD3 supplementation is protective.

*Design.* This study had a paired design comparing individual participants before and after VitD3 supplementation.

*Setting.* Community-based outpatient.

*Patients.* 11 VitD deficient (25-hydroxyvitamin D [25(OH)D] ≤ 20 ng/mL) individuals recruited from the Amish population in Lancaster PA.

*Interventions.* Participants underwent two canagliflozin challenge protocols (300 mg daily for five days): the first before and the second after VitD3 supplementation. In the VitD3 supplementation protocol, participants received VitD3 supplementation (50,000 IU once or twice a week depending on BMI for 4-6 weeks) to achieve 25(OH)D ≥ 30 ng/mL.

*Main Outcome Measures.* Two co-primary endpoints were identified: effects of VitD3 supplementation on canagliflozin-induced changes in 1,25(OH)_2_D and PTH. Secondary endpoints included effects of VitD3 supplementation on baseline levels of VitD metabolites and PTH.

*Results.* VitD3 supplementation increased mean 25(OH)D from 16.5±1.6 to 44.3±5.5 ng/mL (p=0.0006) and 24,25-dihydroxyvitamin D [24,25(OH)_2_D] from 1.0±0.1 to 4.3±0.6 ng/mL (p=0.0002). Mean 1,25(OH)_2_D and PTH were unchanged. VitD3 supplementation decreased the magnitude of canagliflozin-induced changes in 1,25(OH)_2_D (from −31.3%±4.7% to −9.3%±8.3%; p=0.04) and PTH (from +36.2%±6.2% to +9.7%±3.7%; p=0.005).

*Conclusions.* VitD deficiency rendered individuals more vulnerable to adverse effects of canagliflozin on biomarkers associated with bone health. VitD3 supplementation was protective against canagliflozin’s short-term adverse effects on 1,25(OH)_2_D and PTH.

## Introduction

SGLT2 inhibitors display an attractive clinical efficacy profile including glucose-lowering efficacy, decreased risk of major adverse cardiovascular events, decreased risk of hospitalization for heart failure, and slowed progression of kidney disease (1–7). However, there is controversy about the bone related side effects of these drugs. Canagliflozin and dapagliflozin have been reported to increase the risk of bone fracture in some (4, 8) but not all (1, 2, 9) clinical trials. Canagliflozin has also been reported to accelerate loss of bone mineral density (10), which may contribute to the increased risk of bone fracture. We have proposed that adverse effects of SGLT2 inhibitors on bone health are mediated by decreasing levels of 1,25-dihydroxyvitamin D [1,25(OH)_2_D] and increasing levels of PTH (11–14). Canagliflozin, dapagliflozin, and empagliflozin have all been reported to increase serum phosphorus and thereby trigger the FGF23/1,25(OH)_2_D/PTH axis (11, 13, 14), which represent potential endocrine mechanisms that may mediate adverse effects of SGLT2 inhibitors on bone health (12, 15). Although data from most studies assessing the risk of bone fracture in SGLT2 inhibitor users are reassuring, there are important limitations in some of these studies as discussed in the Discussion section.

Vitamin D (VitD) status contributes to determining the state of bone health (16–18). Low 1,25(OH)_2_D and increased levels of PTH work in tandem to negatively impact bone health (19). Moreover, canagliflozin, dapagliflozin, and empagliflozin have all been reported to decrease 1,25(OH)_2_D and increase PTH (11, 13, 14). We hypothesized that low availability of the substrate 25-hydroxyvitamin D [25(OH)D] might render individuals more vulnerable to the canagliflozin-induced decrease in 1α-hydroxylase activity. In other words, VitD deficiency might amplify the canagliflozin-induced changes in 1,25(OH)_2_D and PTH. Accordingly, we investigated potential nutrient-drug interactions between VitD and canagliflozin by conducting a clinical trial in otherwise healthy VitD deficient individuals with baseline total serum level of 25(OH)D ≤ 20 ng/mL in whom we assessed the effects of canagliflozin on 1,25(OH)_2_D, PTH, and FGF23 before and after VitD3 supplementation.

## Materials and Methods

This study (“VitD Sub-study”) was part of our ongoing parent “Genetics of Response to Canagliflozin” clinical trial (clinicaltrials.gov Identifier: NCT02891954) in healthy volunteers recruited from the Old Order Amish population in Lancaster PA, who are non-Hispanic White people. Although participants were excluded from the parent study if their 25(OH)D level was ≤ 20 ng/mL, these VitD deficient individuals were eligible to participate in the VitD Sub-study, which investigated the effects of VitD3 supplements on canagliflozin-triggered changes in the phosphate/FGF23/1,25(OH)_2_D/PTH axis. Participants were required to meet all other exclusion/inclusion criteria for the parent study as listed below.

### Inclusion/exclusion criteria

Inclusion criteria:

- Amish descent
- ≥18 years old
- VitD deficiency at baseline with 25(OH)D ≤ 20 ng/mL
- BMI of 18-40 kg/m^2^
- Participants were required to be willing not to alter their regimen of vitamins, nutritional supplements, and over-the-counter medications (with the exception of acetaminophen) during the course of the study. If changes needed to be made, the PI or designated research physician would review and approve changes in order to remain in the study.

Exclusion criteria:

- Known allergy to canagliflozin or VitD3
- History of diabetes and/or HbA1c > 6.5%
- History of medical condition which, in the judgment of the PI or designated research physician, would alter the response to canagliflozin and/or increase the risk to the patient
- History of low trauma bone fractures associated with a history of osteoporosis
- History of a urinary tract infection or genital yeast infection within the past year
- Current treatment with diuretics, antihypertensive medication, uric acid lowering medications, or other medication that the investigator judges will make interpretation of the results difficult
- Significant debiliatating chronic cardiac, hepatic, pulmonary, or renal disease or other diseases that the investigator judges will make interpretation of the results difficult or increase the risk of participation
- Seizure disorder
- Unhealed foot ulcer
- Pregnant by self-report, positive urine pregnancy test done by nursing staff prior to administration of study drug, or known pregnancy within three months of the start of the study
- Currently breast feeding or breast feeding within three months of the start of the study
- Estimated glomerular filtration rate <60 mL/min/1.73 m^2^
- Liver function tests >2 times the upper limit of normal
- Hematocrit <35%
- Abnormal TSH
- Urinalysis suggesting urinary tract infection.

### Study population

Otherwise, healthy participants were recruited from the Old Order Amish population in Lancaster PA, as described in the Results (“Disposition and baseline characteristics of participants” section).

### Rationale for enrolling individuals with 25(OH)D ≤ 20 ng/mL

The Endocrine Society’s guidelines proposed thresholds of 25(OH)D ≤20 ng/mL (≤ 50 nmol/L) for VitD deficiency, 21-29 ng/mL (51-74 nmol/L) for VitD insufficiency, and ≥ 30 ng/mL (≥ 75 nmol/L) for VitD sufficiency. We applied The Endocrine Society’s definition of VitD deficiency (≤ 20 ng/mL) as a key inclusion criterion and applied the definition of VitD sufficiency (≥ 30 ng/mL) as the criterion for adequacy of VitD3 supplementation.

### Canagliflozin challenge protocol

Research participants underwent two canagliflozin challenges. Canagliflozin challenge #1 was conducted when participants were judged to be VitD deficient [25(OH)D ≤ 20 ng/mL]; canagliflozin challenge #2 was conducted after participants had received VitD3 supplementation and 25(OH)D levels were ≥ 30 ng/mL. Research nurses conducted home visits at approximately 7am to obtain blood samples on Day 1 (pre-dose), Day 3 (after 2 doses of canagliflozin), and Day 6 (after 5 doses of canagliflozin). To minimize the impact of circadian rhythms, blood samples were obtained at the time of these home visits. Canagliflozin (300 mg/d, p.o.) was administered every morning shortly after obtaining 7am blood samples on Days 1-5. Blau et al. (11) previously reported that maximum responses to canagliflozin in a 5-day study were observed after 48 hours (Day 3) for serum phosphorus, FGF23, and 1,25(OH)_2_D and after 120 hours (Day 6) for PTH. These previously published observations (11) provided the rationale for measuring these parameters on Days 1, 3, and 6 and focusing on canagliflozin-triggered changes on Day 6 for PTH, and on Day 3 for the other biomarkers.

### VitD3 supplementation protocol

VitD deficient participants received VitD3 capsules (Bio-Tech Pharmacal; Fayetteville, AR) in doses of either 50,000 IU per week (for participants with BMI <30 kg/m^2^) or 50,000 IU twice a week (for participants with BMI ≥30 kg/m^2^) for four weeks, followed by a second 25(OH)D measurement a week after completion of the four-week VitD3 supplementation protocol. If screening 25(OH)D levels (Quest Diagnostics) were <30 ng/mL, high-dose VitD3 supplementation was continued. Once screening 25(OH)D (Quest Diagnostics) levels were confirmed to be ≥30 ng/mL, maintenance-dose VitD3 supplementation (1000 IU/d or 2000 IU/d for BMI <30 kg/m^2^ or BMI ≥30 kg/m^2^, respectively) was continued at least until completion of canagliflozin challenge #2.

### Clinical chemistry

Blood samples obtained at home visits were collected in test tubes as appropriate for each assay: EDTA anticoagulant (purple top tube) for measurement of plasma samples and red top tube for collecting serum samples. After placing tubes on ice, blood samples were transported to the clinical laboratory at the Amish Research Clinic (maximum transport time, 2 hours). After centrifugation (3300 rpm for 10 min), plasma/serum was sent on the same day to a laboratory for assay.

Most laboratory assays were conducted at Quest Diagnostics except when stated otherwise. Some specialized assays were conducted at the Cytokine Core Laboratory of the University of Maryland School of Medicine using the following kits: intact human FGF23 (iFGF23) and C-terminal FGF23 (cFGF23) (Quidel Corporation, San Diego, CA).

For consistency with the methods of the parent study, screening serum levels of 25(OH)D and all serum levels of 1,25(OH)_2_D were assayed using LC-MS/MS methodology at Quest Diagnostics. As requested by the funding agencies (NIH Office of Dietary Supplements and National Institute of Diabetes and Digestive and Kidney Disease; National Institutes of Health), plasma levels of 25(OH)D and 24,25-dihydroxyvitamin D [24,25(OH)_2_D] were measured using LC-MS/MS methodology by Heartland Assays (Ames, IA). Quest’s 25(OH)D levels were used to determine eligibility for the VitD Sub-study. Heartland’s 25(OH)D levels were used to assess the response to VitD3 supplements. All assays of hydroxylated VitD metabolites provided separate measurements for both VitD2 and VitD3 derivatives. We report 25(OH)D and 24,25(OH)_2_D in ng/mL and 1,25(OH)_2_D in pg/mL. One can use the conversion factor of 2.49 to convert ng/mL to nmol/L for 25(OH)D.

### Co-primary endpoints

We established two co-primary outcomes to investigate whether VitD3 supplements mitigate the canagliflozin-induced changes in biomarkers related to bone health: effect of VitD3 supplements on canagliflozin-induced decrease in 1,25(OH)_2_D and effect of VitD3 supplements on canagliflozin-induced increase in PTH.

### Secondary endpoints

Our secondary endpoints included 2 categories: (a) canagliflozin-dependent endpoints and (b) canagliflozin-independent endpoints.

### Canagliflozin-dependent endpoints

a. Effect of canagliflozin on serum phosphorus (Day 3 versus Day 1) before and after VitD3 supplementation; we compared the canagliflozin-induced changes in serum phosphorus before versus after VitD3 supplementation.
b. Effect of canagliflozin on serum calcium (Day 3 versus Day 1) before and after VitD3 supplementation; we compared the canagliflozin-induced changes in serum calcium before versus after VitD3 supplementation.
c. Effect of canagliflozin on intact FGF23 [iFGF23] (Day 3 versus Day 1) before and after VitD3 supplementation; we compared the canagliflozin-induced changes in iFGF23 before versus after VitD3 supplementation.
d. Effect of canagliflozin on C-terminal FGF23 [cFGF23] (Day 3 versus Day1) before and after VitD3 supplementation; we compared the canagliflozin-induced changes in cFGF23 before versus after VitD3 supplementation.

We used paired t-tests for all analyses.

### Canagliflozin-independent endpoints

a. We compared baseline values of plasma 1,25(OH)_2_D and 24,25(OH)_2_D before versus after VitD3 supplementation, independent of canagliflozin.
b. We compared baseline values of serum phosphorus, calcium, plasma PTH, iFGF23, and cFGF23, before versus after VitD3 supplementation, independent of canagliflozin.

### Statistical analysis

We applied a stepwise approach to test statistical significance for our primary endpoints:

1. Paired t-test to confirm that VitD3 supplements increased levels of 25(OH)D.
2. Paired t-test to confirm that canagliflozin decreased levels of 1,25(OH)_2_D in VitD deficient patients. This comparison was based on ratios of 1,25(OH)_2_D levels on Day 3:Day 1.
3. Paired t-test to investigate whether VitD3 supplements affected the magnitude of canagliflozin-induced decrease in levels of 1,25(OH)_2_D. This assessment was based on a comparison of ratios of 1,25(OH)_2_D levels on Day 3:Day 1 in VitD deficient patients versus the same ratio in patients who had received VitD3 supplements.
4. Paired t-test to confirm that canagliflozin increased levels of PTH in VitD deficient patients. This comparison was based on ratios of PTH levels on Day 6:Day 1.
5. Paired t-test to investigate whether VitD3 supplements affected the magnitude of canagliflozin-induced increase in levels of PTH. This assessment was based on a comparison of ratios of PTH levels on Day 6:Day 1 in VitD deficient patients versus the same ratio in patients who had received VitD3 supplements.

These statistical tests of significance were conducted in sequential fashion. If a statistical analysis yielded a statistically significant result (p-value ≤ 0.05), then we were permitted to proceed to the next comparison. If a statistical analysis did not yield a statistically significant result (p-value > 0.05), then the subsequent analyses would be viewed as exploratory. A two-sided p-value ≤ 0.05 (paired t-tests) was taken as the threshold for statistical significance without any correction for multiple comparisons. For two analysesu, we pooled all the data before and after VitD3 supplementation; in those two analyses, we calculated values for Pearson or Spearman correlation coefficients (Excel).

### Power calculations

We did not conduct a prospective power calculation before designing this study as we did not know whether VitD3 supplementation would affect the magnitude of canagliflozin-induced changes in our primary endpoints [PTH and 1,25(OH)_2_D] nor did we know the magnitude of the variances in responses to VitD3 supplementation. Now that data are available, we performed a retrospective power calculation using data from this study. Based on those assumptions, we calculated that a sample size of N=8 would provide 80% power to detect a significant effect of VitD3 supplements with an α=0.05 (paired t-test). We conducted a similar power calculation for 1,25(OH)_2_D, which estimated that a sample size of N=18 would be required to achieve 80% power with an α=0.05 (paired t-test). Accordingly, we conclude in retrospect that our sample size of N=11 provided >80% power to detect a significant effect of VitD3 supplementation on the canagliflozin-induced change in PTH but <80% power to detect a significant effect of VitD3 supplementation on the canagliflozin-induced change in 1,25(OH)_2_D.

### Study approval

The Clinical Trial Protocol was approved by the University of Maryland Baltimore Institutional Review Board (FWA00007145). Informed consent was obtained from all participants.

## Results

### Disposition and baseline characteristics of participants

Research participants (N=11) were recruited from the Old Order Amish population in Lancaster PA. The disposition of participants is summarized in a CONSORT diagram (Fig. 1). The parent study (“Genetics of the Response to Canagliflozin”) identified 24 individuals with 25(OH)D levels ≤ 20 ng/mL between September, 2019 and July, 2021 who were assessed for eligibility for this VitD sub-study. Nineteen participants gave informed consent and were enrolled in the study. Three participants withdrew consent and 16 completed the first canagliflozin challenge test. Two participants were subsequently withdrawn because they did not achieve 25(OH)D levels ≥ 30 ng/mL in response to VitD3 supplements. Three participants withdrew because of interruptions associated with the COVID-19 pandemic. Eleven participants (7 males and 4 females) completed the entire study. Baseline characteristics and demographics are shown in Table 1. Our study population had a mean age of 48.4 ± 3.2 years and a mean BMI of 29.0 ± 1.2 kg/m^2^ at baseline.

**Figure 1.**
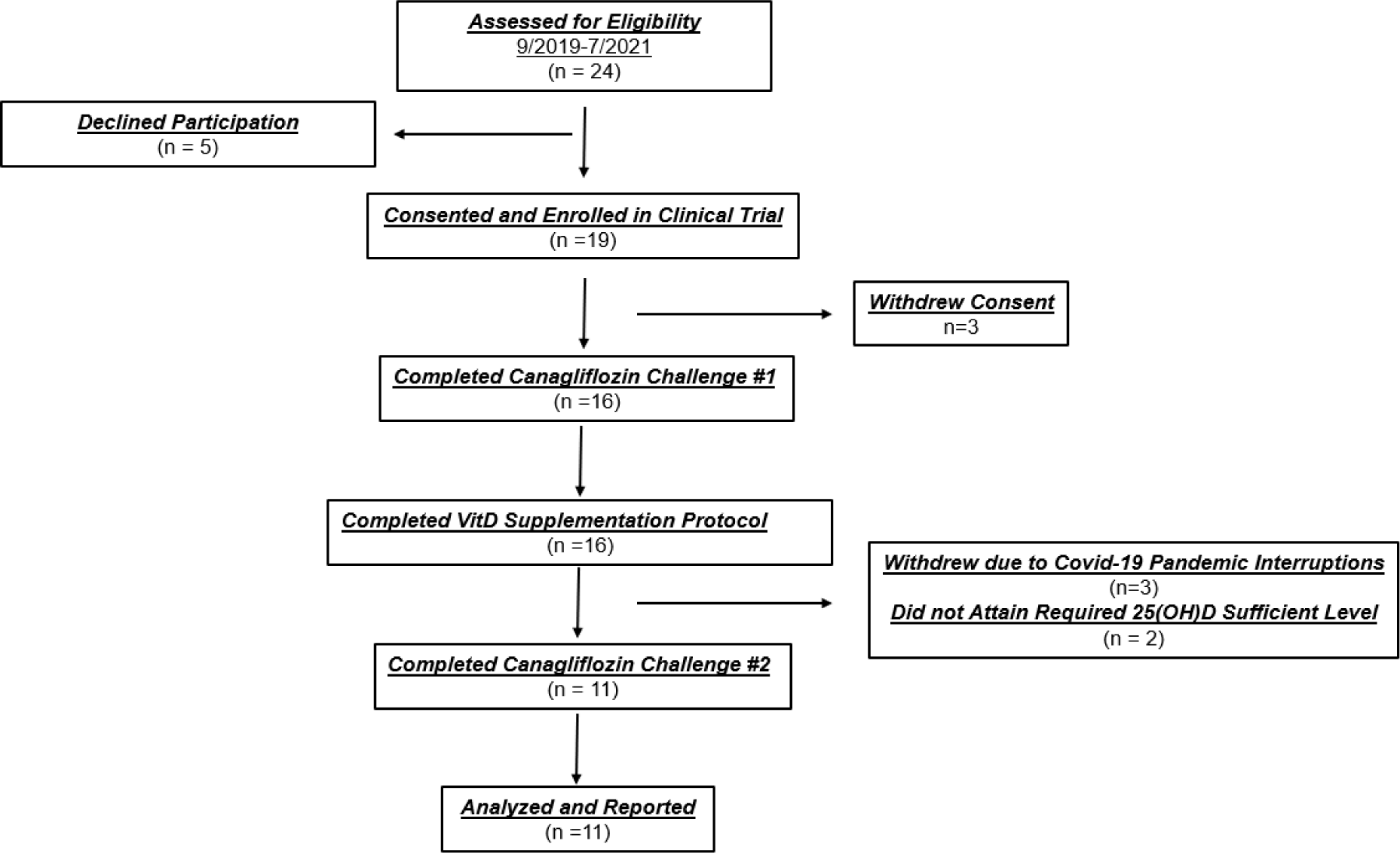
CONSORT diagram. The figure summarizes recruitment and disposition of research participants in this clinical trial.

**Table 1.** Participants’ demographics and baseline characteristics (mean ± SEM).

|  | **Total** | **Females** | **Males** |
| --- | --- | --- | --- |
| N | 11 | 4 | 7 |
| Age (years) | 48.0 ± 3.2 | 49 ± 5.3 | 48 ± 4.4 |
| Height (cm) | 170 ± 2.4 | 163.3 ± 2.0 | 173.9 ± 2.6 |
| Weight (kg) | 81.7 ± 3.1 | 77.5 ± 5.4 | 84.5 ± 3.6 |
| BMI (kg/m<sup>2</sup>) | 29.0 ± 1.2 | 28.8 ± 1.5 | 28.9 ± 1.7 |
| HbA1c (%) | 5.4 ± 0.08 | 5.5 ± 0.1 | 5.3 ± 3.08 |
| eGFR (mL/min/1.73 m<sup>2</sup>) | 98.5 ± 4.0 | 100 ± 8.6 | 98.6 ± 3.5 |

### Effect of VitD3 supplementation on VitD metabolites and mineral metabolism biomarkers

Clinical trial participants were required to have baseline 25(OH)D levels ≤ 20 ng/mL at the time of screening. For consistency with the methods in the parent study (“Genetics of Response to Canagliflozin”), we used a 25(OH)D assay provided by Quest Diagnostics to screen people for eligibility for both the parent study and the VitD sub-study. To maximize the ability to compare with data on 24,25(OH)_2_D levels, Heartland Assays provided assays for both 25(OH)D and 24,25(OH)_2_D to analyze blood samples collected during the canagliflozin challenge tests conducted as part of the VitD sub-study. Figure 2 demonstrates that the two assays (i.e., Quest Diagnostics and Heartland Assays) yielded essentially identical results for mean levels of 25(OH)D and exhibited good overall correlation for individual participants (r=0.9; p=10^-8^). Blood samples for screening were collected in advance of (typically one or two weeks before) the canagliflozin challenge tests. The comparability between the two sets of data is reassuring given that the samples were not collected on the same day and the assays were conducted in different laboratories.

**Figure 2.**
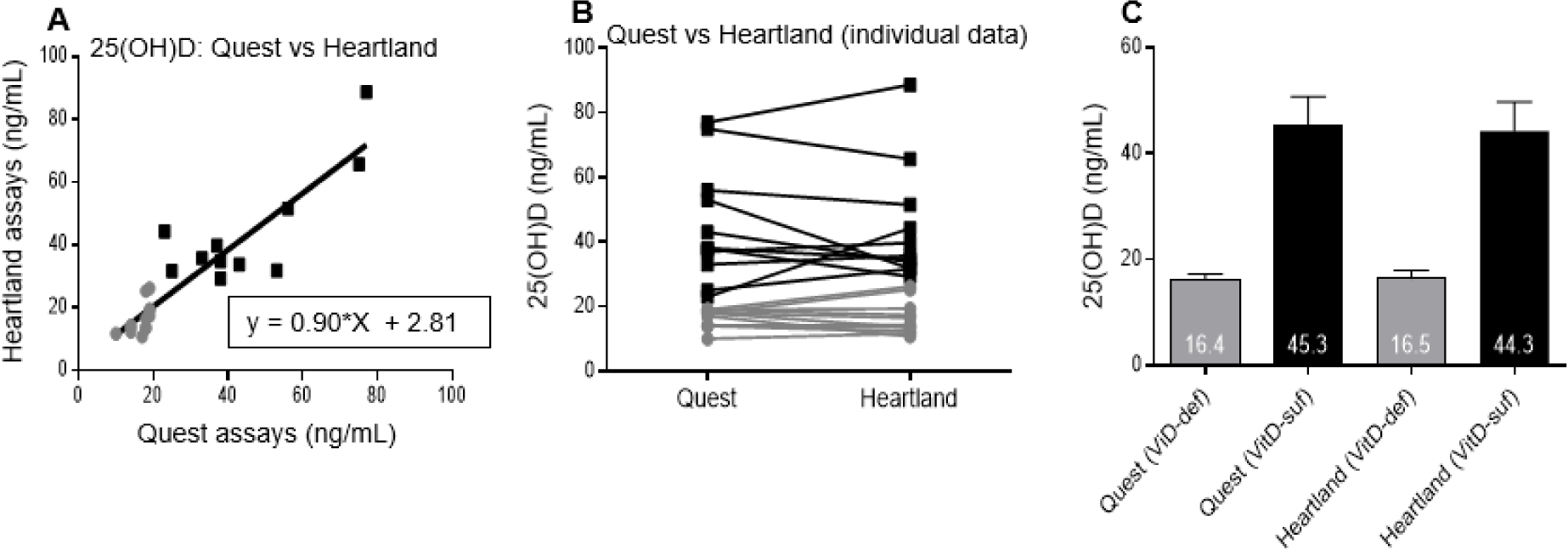
Comparison of two commercial assays for 25(OH)D in participants in our clinical trial: Quest Diagnostics and Heartland Assays. Panel A plots individual levels of total 25(OH)D measured by Heartland Assays versus individual levels of total 25(OH)D measured by Quest Diagnostics. Panel B compares measurements of 25(OH)D levels in the two assays at the level of individual participants when they were VitD deficient (gray circles) and after they received VitD3 supplements (black squares). Panel C shows means ± SEM for total 25(OH)D levels in VitD-deficient participants before (gray bars) and after receiving VitD3 supplements (black bars) as measured by Quest Diagnostics and Heartland Assays.

Supplementation with VitD3 significantly increased mean (± SEM) 25(OH)D by 2.7-fold from 16.5 ± 1.6 ng/mL in the VitD deficient state to 44.3 ± 5.5 ng/mL after VitD3 supplementation (p=0.0006) (Fig. 3A). Mean 24,25(OH)_2_D levels increased 4.3-fold from 1.0 ± 0.1 ng/mL in the VitD deficient state to 4.3 ± 0.6 ng/mL after VitD3 supplementation (p=0.0002) (Fig. 3B). In contrast, mean plasma levels of 1,25(OH)_2_D in the VitD deficient state did not change significantly after VitD3 supplementation (43.8 ± 3.6 versus 44.9 ± 4.1 pg/mL; p=0.7) (Fig. 3C).

**Figure 3.**
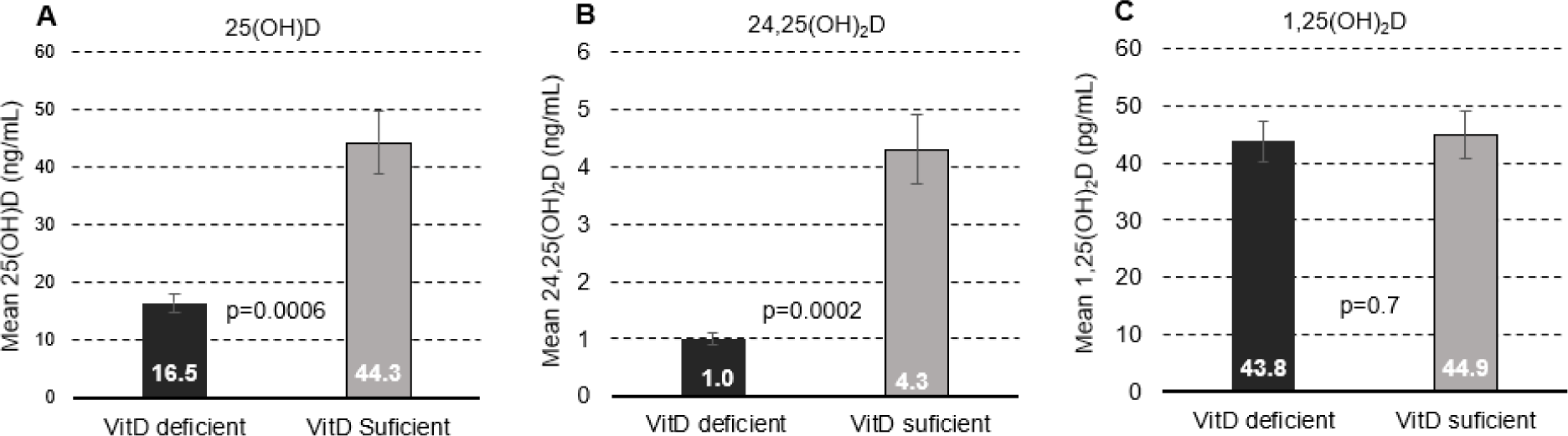
Effect of VitD3 supplementation on mean baseline plasma levels of hydroxylated VitD metabolites: 25(OH)D (panel A), 24,25 (OH)_2_D (panel B), and 1,25(OH)_2_D (panel C). Mean (±SEM) baseline plasma levels of VitD metabolites were measured when research participants (N=11) were VitD deficient (black bars) and again after participants received VitD3 supplements (gray bars). Mean levels of VitD metabolites are indicated in white font. Error bars represent standard errors of the mean. Levels of statistical significance are indicated by p-values using Student’s t-test for paired comparisons.

### Mean levels of plasma PTH and FGF23 did not change in response to VitD3 supplementation

Mean levels of serum phosphorus, serum calcium, plasma PTH, and plasma FGF23 (intact and C-terminal) were within the normal range when participants were VitD deficient and did not change with VitD3 supplementation (Table 2). 25(OH)D levels spanned an 8-fold range (11 - 88 ng/mL); PTH levels ranged between 22-85 pg/mL (Fig. 4A) and were not significantly correlated with 25(OH)D levels in an analysis of our pooled data (Fig. 4A). Furthermore, we did not observe a consistent effect of VitD3 supplementation on PTH levels of individual participants (Fig. 4B). Baseline levels of PTH were closely correlated with PTH levels after administration of VitD3 supplements (r=0.83; p=0.001) (Fig. 4C). Although the published literature provides extensive evidence that VitD deficiency can cause secondary hyperparathyroidism, It is suggested that secondary hyperparathyroidism is most commonly triggered in the context of more severe VitD deficiency [e.g., total 25(OH)D levels <10-12 ng/mL] (20–23).

**Figure 4.**
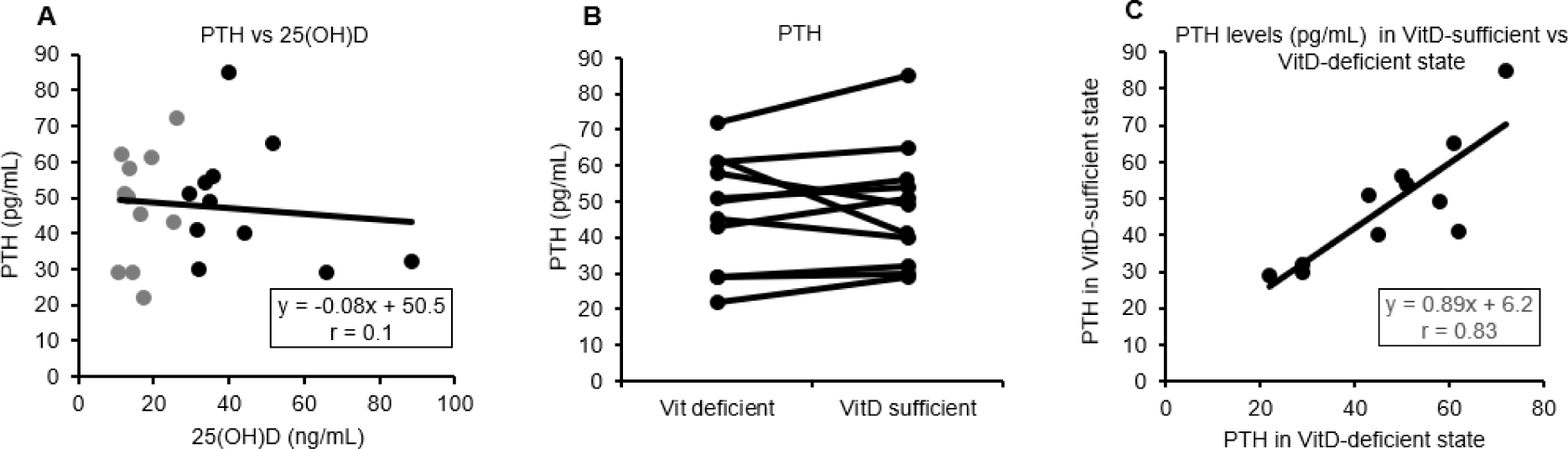
Effect of VitD status on PTH levels. Panel A presents pooled data for individual participants when they were VitD deficient (gray circles) and after they received VitD3 supplements (black circles). PTH levels are plotted as a function of total plasma levels of 25(OH)D. Panel B plots levels of PTH when individual participants were VitD deficient versus plasma levels of PTH after they received VitD3 supplements. Panel C plots PTH levels after participants received VitD3 supplements as a function of PTH levels when participants were VitD deficient. The text boxes in panels A and C present the equations for the linear regression lines and the Pearson correlation coefficients.

**Table 2.** Effect of VitD3 supplementation on levels of serum phosphorus, serum calcium, plasma PTH, plasma iFGF23, and cFGF23. The Table summarizes mean levels (±SEM) when participants (N=11) were VitD deficient and after they received VitD3 supplements. p-values were calculated using Student’s t-test for paired comparisons.

|  | VitD deficient state | VitD sufficient state | p-values |
| --- | --- | --- | --- |
| Serum Phosphorus (mg/mL) | 3.6 ± 0.1 | 3.7 ± 0.1 | 0.3 |
| Serum Calcium (mg/mL) | 9.3 ± 0.1 | 9.2 ± 0.1 | 0.4 |
| Plasma PTH (pg/mL) | 47.5 ± 4.8 | 48.4 ± 5.1 | 0.8 |
| Plasma iFGF23 (pg/mL) | 43.1 ± 5.6 | 46.5 ± 5.0 | 0.2 |
| Plasma cFGF23 (RU/mL) | 76.5 ± 13.2 | 83.0 ± 14.0 | 0.2 |

### Interaction between baseline VitD status and pharmacodynamic responses to canagliflozin

When participants were VitD deficient, canagliflozin significantly decreased mean (± SEM) 1,25(OH)_2_D levels by 31.3% ± 4.7%: from 43.8 ± 3.6 pg/mL on Day 1 to 29.1 ± 2.3 pg/mL on Day 3 (p=0.0003). In contrast, after participants had received VitD3 supplements, canagliflozin induced a smaller, non-significant 9.3% ± 8.3% decrease in mean levels of 1,25(OH)_2_D: from 45 ± 4.1 pg/mL on Day 1 to 41 ± 5.4 pg/mL on Day 3 (p=0.3). Thus, VitD3 supplementation provided statistically significant protection from the adverse effect of canagliflozin to decrease mean plasma levels of 1,25(OH)_2_D (p=0.04) (Fig. 5A). Similarly, when participants were VitD deficient, canagliflozin triggered a significant 36.2% ± 6.2% increase in mean levels of PTH: from 47.5 ± 4.8 pg/mL on Day 1 to 58.5 ± 5.4 pg/mL on Day 6 (0.0009). After participants had received VitD3 supplements, canagliflozin induced a smaller mean increase (9.7% ± 3.7%) in mean plasma levels of PTH: from 48.4 ± 5.1 pg/mL on Day 1 to 53.3 ± 6.1 pg/mL on Day 6 (p=0.02). Therefore, VitD3 supplementation significantly decreased the magnitude of the canagliflozin-induced increase in mean levels of PTH (p=0.005) (Fig. 5B).

**Figure 5.**
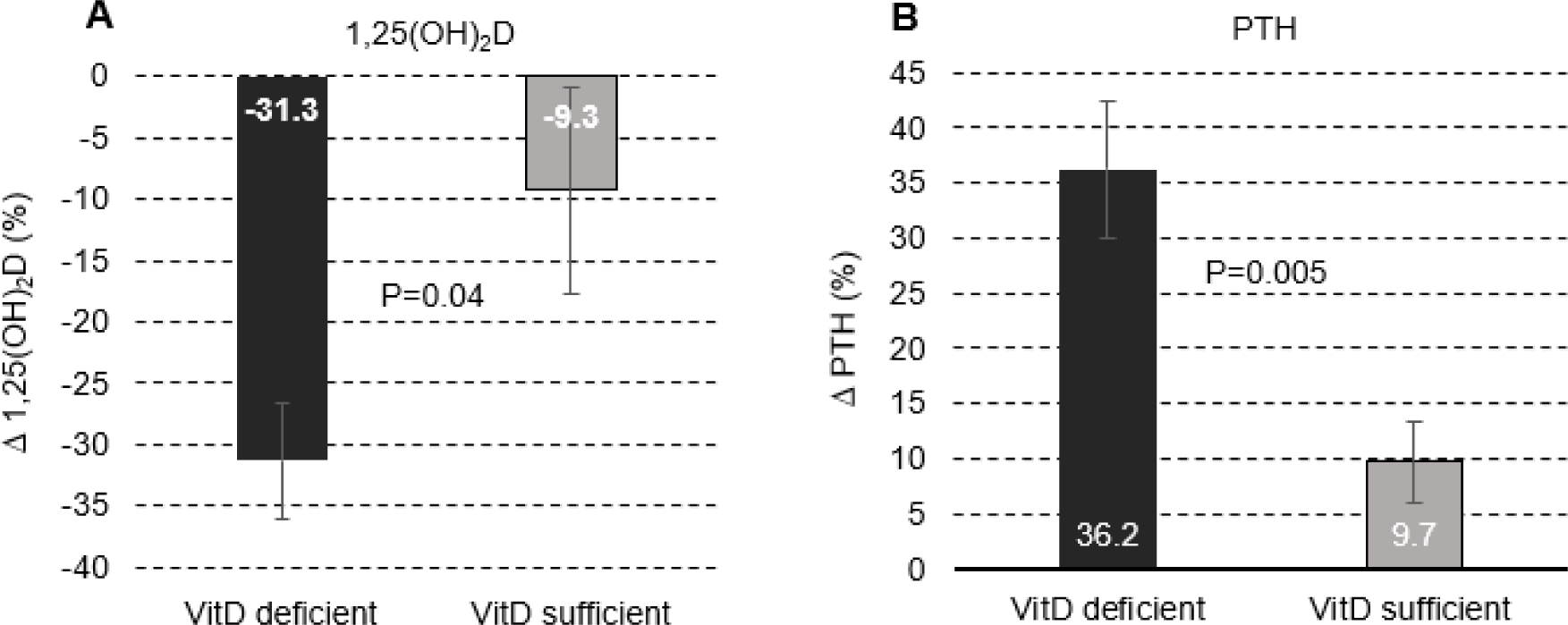
Canagliflozin-induced change in primary endpoints: plasma 1,25(OH)_2_D and PTH. Research participants (N=11) underwent two canagliflozin challenge protocols: the first when they were VitD deficient (black bars) and the second after receiving VitD supplements (gray bars). Mean canagliflozin-induced percentage changes (depicted in white font) are shown for Day 6 for PTH and for Day 3 for 1,25(OH)_2_D. Error bars represent standard errors of the mean. p-values were calculated using Student’s t-test for paired comparisons for the effects of VitD.

### Effect of VitD on canagliflozin-induced changes in secondary endpoints

In the VitD deficient state, canagliflozin significantly increased mean (± SEM) serum phosphorus on Day 3 versus Day 1 (3.6 ± 0.1 mg/dL to 3.8 ± 0.1 mg/dL; p=0.007) with the mean percentage change of +7.0% ± 2.1% (Table 3). In contrast, the percentage change was −0.2% ± 3.8% after VitD3 supplementation (3.72 ± 0.1 mg/dL to 3.69 ± 0.2 mg/dL; p=0.8). While the canagliflozin-induced change in serum phosphorus was numerically larger in the VitD deficient state, the impact of VitD3 supplementation did not achieve statistical significance (p=0.1).

**Table 3.**
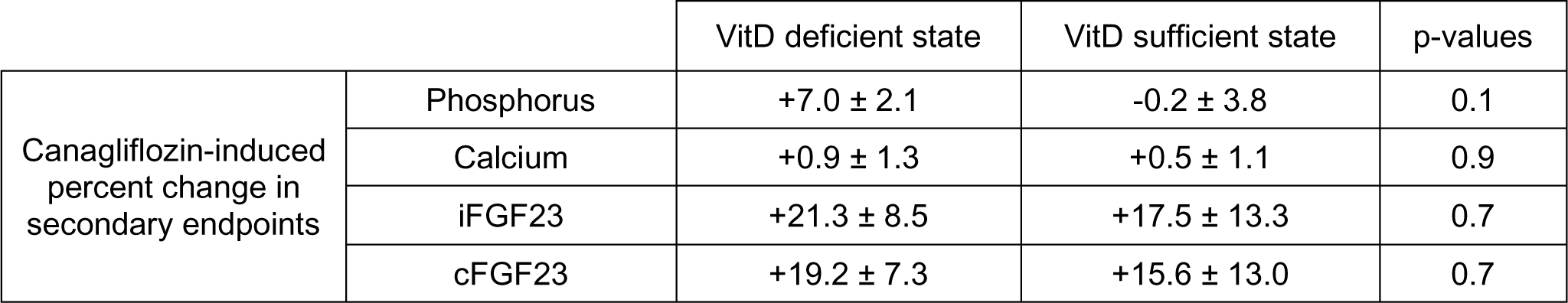
Effects of VitD3 supplementation on canagliflozin-induced changes in secondary endpoints. The Table summarizes canagliflozin-induced changes (mean ± SEM) in serum phosphorus, serum calcium, plasma iFGF23, and cFGF23. p-values were calculated using Student’s t-test for paired comparisons for the effects of canagliflozin.

Canagliflozin increased iFGF23 by 21.3% when participants were VitD deficient (from 43.1 ± 5.6 pg/mL on Day 1 to 50.9 ± 6.2 pg/mL on Day 3; p=0.009) and by 17.4% after VitD3 supplementation (from 46.5 ± 5.0 pg/mL on Day 1 to 54.2 ± 8.9 pg/mL on Day 3; p=0.2) (Table 3). Canagliflozin also increased cFGF23 by 19.2% (from 76.5 ± 13.2 RU/mL on Day 1 to 86.4 ± 13.2 RU/mL on Day 3; p=0.08) before VitD3 supplementation and by 15.6% (from 83.0 ± 14.0 RU/mL on Day 1 to 94.3 ± 20.8 RU/mL on Day 3; p=0.3) after VitD3 supplementation (Table 3). 1,25(OH)_2_D has been reported to increase the expression of the *FGF23* gene (24). It is possible, therefore, to envision interactions between VitD and the magnitude of canagliflozin’s effect on iFGF23 even though we did not observe a statistically significant effect of VitD3 supplementation on the magnitude of the canagliflozin-induced increase in either iFGF23 or cFGF23 – possibly because the sample size was not sufficiently large.

### Effect of canagliflozin on C-terminal telopeptide of type 1 collagen (CTX) and procollagen type 1 N-terminal propeptide (P1NP)

When participants were VitD deficient, canagliflozin administration increased CTX from 389 ± 40 on Day 1 to 455 ± 66 pg/mL on Day 6 (Fig. 6A). This 17% increase had a p-value of 0.058, which was close to the threshold for statistical significance. In contrast, CTX levels did not change significantly in response to canagliflozin after participants had received VitD3 supplementation (430 ± 61 to 454 ± 59 pg/mL; p= 0.39) (Fig. 6B). Canagliflozin administration was associated with a numerical trend toward a decrease of P1NP by 7.4% (33.6 ± 3.1 to 31.1 ± 4.3 ng/mL; p= 0.21) in the VitD deficient state (Fig. 7A). Canagliflozin had no significant effect on P1NP after VitD3 supplementation (36.4 ± 3.1 versus 35.6 ± 3.6 ng/mL; p= 0.67) (Fig. 7B).

**Figure 6.**
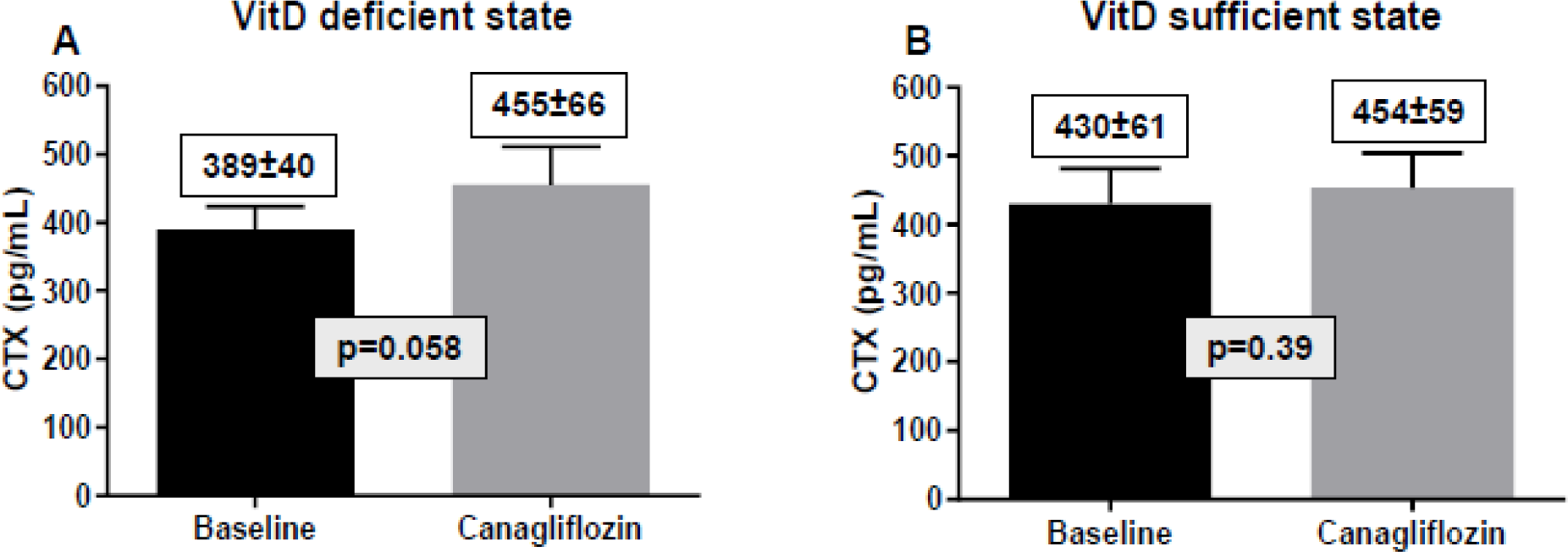
Effects of canagliflozin on a bone resorption biomarker (C-terminal telopeptide of type 1 collagen (CTX)). Panel A illustrates the mean (±SEM) level of CTX on Day 1 (black bar) and after 5 days of treatment with canagliflozin (gray bar) when participants were VitD deficient. Panel B illustrates the mean (±SEM) level of CTX on Day 1 (black bar) and after 5 days of treatment with canagliflozin (gray bar) when participants were VitD sufficient.

**Figure 7.**
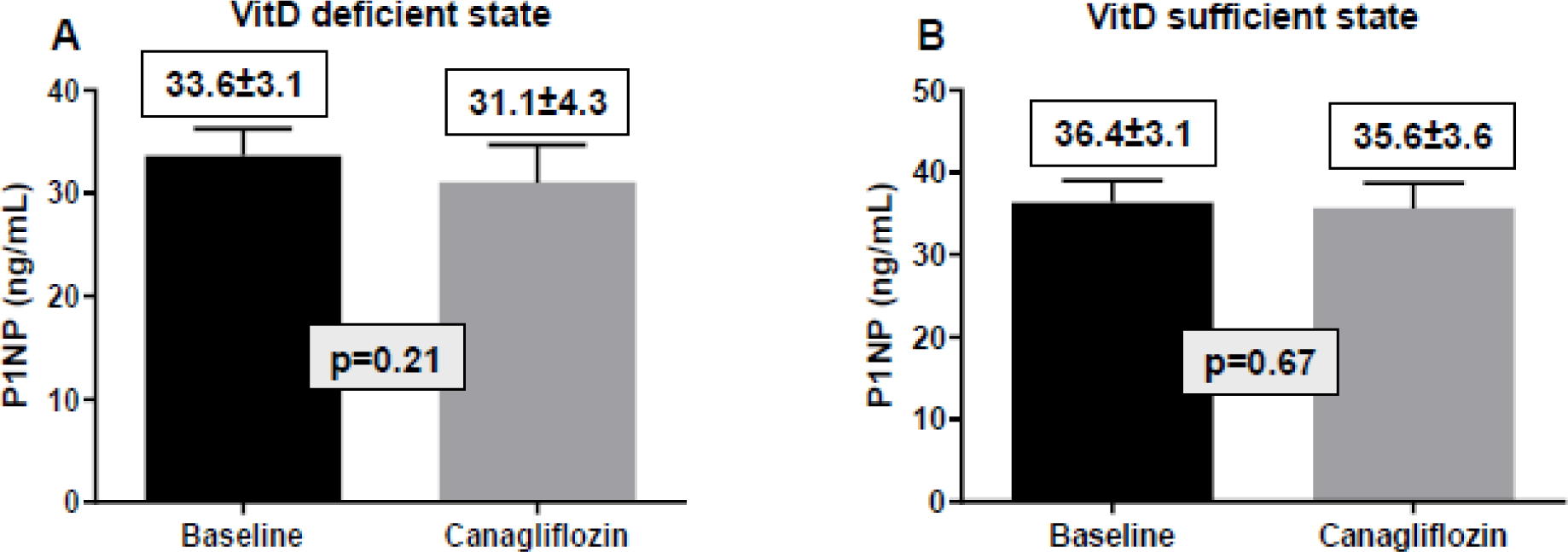
Effects of canagliflozin on a bone formation biomarker (Procollagen type 1 N- terminal propeptide (P1NP)). Panel A illustrates the mean (±SEM) level of P1NP on Day 1 (black bar) and after 5 days of treatment with canagliflozin (gray bar) when participants were VitD deficient. Panel B illustrates the mean (±SEM) level of P1NP on Day 1 (black bar) and after 5 days of treatment with canagliflozin (gray bar) when participants were VitD sufficient.

## Discussion

Canagliflozin inhibits SGLT2-mediated sodium and glucose co-transport in the proximal tubule. This increases the electrochemical gradient for Na^+^, which in turn increases the drive for sodium-dependent reabsorption of phosphate. The resulting increase in serum phosphorus is associated with an increase in plasma FGF23, which in turn decreases expression of 1α-hydroxylase in the kidney and decreases plasma levels of 1,25(OH)_2_D. This would decrease intestinal absorption of dietary calcium, which ultimately increases PTH secretion (11, 12, 15). We have previously reported that canagliflozin decreased 24-hour urinary excretion of calcium and phosphate (11). The decrease in urinary phosphate excretion was likely driven by the observed increases in FGF23 and PTH. The decrease in urinary calcium excretion was likely driven by either a decrease in intestinal calcium absorption and/or the observed increase in PTH.

Low levels of 1,25(OH)_2_D and secondary hyperparathyroidism work in tandem to adversely impact bone health (19). Based on our previous observations that five days of treatment with canagliflozin decreased 1α-hydroxylation of 25(OH)D and increased PTH, we hypothesized that low levels of 25(OH)D [the biosynthetic precursor of 1,25(OH)_2_D] would increase vulnerability to the pathophysiological impact of canagliflozin on 1α-hydroxylation. To test our hypothesis, we conducted a five-day clinical physiology study with biomarkers as endpoints. We reproduced our previous observations that canagliflozin adversely impacted 1,25(OH)_2_D and PTH (11). Furthermore, the present study demonstrated that VitD status affects the magnitude of the body’s response to canagliflozin and that VitD3 supplementation mitigated those canagliflozin-induced adverse changes on 1,25(OH)_2_D and PTH (Fig. 5).

Several lines of evidence suggest that SGLT2 inhibitors exert adverse effects on bone health. Canagliflozin and dapagliflozin have been reported to increase the risk of treatment-emergent bone fractures (4, 8). Canagliflozin has also been reported to accelerate the loss of bone mineral density (10). Furthermore, canagliflozin, dapagliflozin, and empagliflozin have been reported to trigger the phosphate/FGF23/1,25(OH)_2_D/PTH axis, which represents a biologically plausible mechanism whereby SGLT2 inhibitors might exert adverse effect on bone health (12–14). On the other hand, several studies did not observe an increased risk of bone fractures (1–3, 25–28). Although it is not clear how to explain this discrepancy, it is possible that differences in patient populations might contribute. Our current clinical trial demonstrated that VitD deficiency increased vulnerability of participants to the adverse effects of canagliflozin on pharmacodynamic biomarkers – specifically 1,25(OH)_2_D and PTH. It is possible that differences in VitD status might have contributed to discrepant outcomes among clinical trials with respect to the risk of bone fracture and may help to explain why treatment with SGLT2 inhibitors was associated with increased risk of bone fractures in some studies but not in others. Interestingly, mean baseline plasma 25(OH)D levels were relatively low in the studies demonstrating that dapagliflozin and empagliflozin trigger decreased levels of 1,25(OH)_2_D and increased levels of PTH. Specifically, 25(OH)D levels were 18 ng/mL in the study with dapagliflozin (13) and 15.6 ng/mL in the study with empagliflozin (14). It is also possible that patients’ baseline bone mineral densities differed among the clinical trials. For example, patients with higher baseline bone mineral density would be predicted to have longer lag times before they lose sufficient bone mineral density to be at risk for fragility fractures. Finally, follow-up times varied among the studies (4, 29–32). As shown in studies with other drugs (e.g., thiazolidinediones), long lag times may be required before an increase in the risk of fracture becomes apparent (33). Some studies (especially studies of “real world” experience) were relatively short (e.g., one year of follow-up) (32); it is likely that longer follow-up might be required to detect an increased risk of bone fracture.

### Study design: strengths and limitations

To maximize statistical power, the study was designed as a crossover study so that we could conduct paired statistical analyses comparing data for individual participants when they were VitD deficient to data obtained after participants received VitD3 supplements. Because our principal endpoints [levels of serum phosphorus, plasma FGF23, plasma 1,25(OH)_2_D, and PTH] all demonstrate circadian rhythms (11, 34, 35), our study design assured that all blood samples would be obtained at the same time of day (∼7:00 am) to minimize the impact of variation due to circadian rhythms. Our LC-MS/MS-based assays of VitD metabolites (provided by Quest and Heartland) distinguished between VitD2 and VitD3. We did not detect metabolites of VitD2 in any of our participants. The absence of VitD2 metabolites avoided potential issues related to differences between VitD2 and VitD3 (36), which simplified data interpretation. Our preliminary data guided us to obtain blood samples at the times when canagliflozin exerted maximal effects on the study’s endpoints (11): Day 6 for PTH and Day 3 for serum phosphorus, FGF23, and 1,25(OH)_2_D. Furthermore, we obtained data on levels of 25(OH)D and 24,25(OH)_2_D from a company (Heartland Assays) that specializes in applying liquid chromatography and mass spectroscopy (LC-MS/MS) to conduct scientifically rigorous and reproducible assays of VitD metabolites.

In addition to these strengths of our study design, the study had important limitations. Although our original study design proposed to study 25 participants, the impact of the COVID-19 pandemic necessitated a decrease in the sample size to 11 participants. As discussed in the Methods, we conducted a retrospective power calculation assuming that VitD3 supplementation decreased the magnitude of the canagliflozin-induced change in PTH by 26.5% with a SD of 22.4% and decreased the magnitude of the canagliflozin-induced change in 1,25(OH)_2_D by 22% with an SD of 31%. Accordingly, we conclude in retrospect that our sample size of N=11 provided >80% power to detect a significant effect of VitD3 supplementation on the canagliflozin-induced change in PTH. Indeed, we observed a significant effect (p=0.005; paired t-test) of VitD3 supplementation to decrease the canagliflozin-induced change in PTH. While our sample size of N=11 provided <80% power to detect an effect on our 1,25(OH)_2_D endpoint, we nevertheless observed a significant effect of VitD3 supplements to decrease the magnitude of the canagliflozin-induced effects on 1,25(OH)_2_D levels (p=0.04; paired t-test).

The study’s endpoints were all based on biomarkers rather than hard clinical outcomes (e.g., fractures). However, the near-significant trend (p=0.058) toward an increase in CTX when participants were VitD deficient suggests an increase in bone resorption which is believed to be predictive of an increase in bone fracture risk (37). We hypothesize that this trend might have achieved statistical significance if the duration of canagliflozin treatment had been longer or if the sample size had been larger. Since none of our participants exhibited 25(OH)D levels below 10 ng/mL, our analyses were limited to individuals with mild or moderate degrees of VitD deficiency.

Limitations associated with the relatively small sample size are partially mitigated by our efforts to assure scientific rigor and reproducibility of the primary data. The crossover study design increased statistical power by enabling paired statistical comparisons. As a result, many of our analyses [e.g., VitD-induced changes in 25(OH)D and 24,25(OH)_2_D] demonstrated high levels of statistical significance. Other analyses [e.g., comparisons for 1,25(OH)_2_D and PTH] demonstrated essentially identical mean values before and after VitD3 supplementation.

This study was designed as an outpatient study with participants free to eat *ad libitum*. Although potential day-to-day variation in dietary intake of calcium, phosphate, and sodium may have increased variation in measured endpoints, the outpatient setting is more representative of “real world” conditions. As part of the parent study (Genetics of Response to Canagliflozin), we collected a 24-hour urine on Day 3 of each canagliflozin challenge test to measure canagliflozin-induced glucosuria. However, we did not obtain baseline 24-hour urine collections prior to administration of canagliflozin, which would have been required to assess the effect of canagliflozin on urinary excretion of calcium or phosphate in this study. Rather, as discussed above, we relied on observations from our previously published in-patient study conducted in a metabolic ward with a fixed metabolic diet (11).

### Effect of VitD3 supplementation on canagliflozin-independent endpoints

VitD3 supplementation increased mean total plasma 25(OH)D and 24,25(OH)_2_D levels but did not change mean 1,25(OH)_2_D and PTH levels. As documented in the literature, the active form of VitD – 1,25(OH)_2_D – is maintained in a physiological range even in the face of mild-moderate VitD deficiency (38). Thus, clinical guidelines recommend against measuring 1,25(OH)_2_D to assess VitD status (20). Similarly, secondary hyperparathyroidism tends to occur mostly in the context of more severe VitD deficiency (e.g., 25(OH)D < 10-12 ng/mL) (21, 22, 39). Inspired by evidence from the literature and our present findings, we developed a mathematical model to investigate homeostatic mechanisms that maintain 1,25(OH)_2_D and PTH in a relatively narrow physiological range (40). Our mathematical model suggests that low availability of VitD suppresses 24- hydroxylation (40), which in turn decreases the rate at which 1,25(OH)_2_D is degraded. Based on our results, we propose that suppression of 24-hydroxylation provides a first line of defense to maintain physiological levels of 1,25(OH)_2_D notwithstanding a mild-moderate deficiency of 25(OH)D (40). This model of homeostatic regulation provides insights into our observations with canagliflozin. Canagliflozin decreases the rate at which 1,25(OH)_2_D is produced as evidenced by the observed decrease in plasma 1,25(OH)2D levels induced by canagliflozin Because the *CYP24A1* gene is regulated by 1,25(OH)_2_D (41), the canagliflozin-induced decrease in 1,25(OH)_2_D would be expected to decrease expression of 24-hydroxylase, which in turn would tend to restore 1,25(OH)_2_D levels toward normal. However, based on the results from our mathematical modeling, we hypothesize that VitD deficiency maximally (80-90%) suppresses 24- hydroxylase activity (40), which prevents further suppression of 24-hydroxylase in response to canagliflozin. In contrast, VitD3 supplementation induces high expression of 24-hydroxylase. These high levels of 24-hydroxylase create a window for 24-hydroxylase to be suppressed in response to canagliflozin-induced impairment of 1,25(OH)_2_D production. Thus, VitD supplementation restores the ability of the body to compensate for the adverse effects of canagliflozin on the phosphorus/FGF23/1,25(OH)_2_D/PTH axis (11).

### Clinical implications

Bilezikian et al. recommended against a one-size-fits-all strategy for assessment of VitD status and management of VitD dosing (42). Rather, they recommended a “tailored” therapeutic approach based on the operative pathophysiological mechanisms in each patient. SGLT2 inhibitors are typically administered to patients with various diseases associated with an increased risk of bone fracture: type 2 diabetes, chronic kidney disease, and/or post-menopausal osteoporosis (1, 42–46). In this clinical setting, patients would be predicted to be especially vulnerable to a “second hit” on bone health – for example, effects of SGLT2 inhibitors to decrease levels of 1,25(OH)_2_D levels and increase levels of PTH. This reasoning suggests that it may be appropriate for physicians to implement an enhanced screening strategy for VitD deficiency in patients receiving SGLT2 inhibitors. Alternatively, in light of the relative safety and low cost of over the counter VitD supplements, it may be appropriate to consider prescribing VitD supplements to all patients receiving SGLT2 inhibitors. Such a strategy might possibly be more cost-effective than a strategy requiring frequent biochemical screening for VitD deficiency.

From a purely scientific perspective, it would be ideal to conduct a large multi-center trial evaluating the impact of VitD supplementation on the incidence of fractures in SGLT2 inhibitor-treated patients. However, it seems quite unlikely that funding will be available to conduct a large and expensive clinical trial to provide a rigorous test of the hypothesis that VitD supplements would protect against adverse effects of SGLT2 inhibitors on bone health. Also, such a study would raise challenging ethical questions related to whether it is ethical to withhold VitD supplements for long periods of time in this clinical setting. Nevertheless, there would be value in conducting a shorter clinical trial (e.g., ∼6 months) studying interactions between VitD3 supplements and SGLT2 inhibitors in patients with type 2 diabetes and/or chronic kidney disease. Even if ethical and financial considerations prevented powering a clinical trial to assess the impact of SGLT2 inhibitors on incidence of bone fractures, it would be informative to study other endpoints – e.g., biomarkers such as 1,25(OH)_2_D, PTH, β-CTX, P1NP, and bone mineral density. We hope our data provide a rationale for funding agencies to invest in future research to address this important clinical question.

## Acknowledgements

The authors gratefully acknowledge research funding provided by NIDDK and the NIH Office of Dietary Supplements: R01DK118942, R01DK118942-02S1, and T32DK098107. We are also grateful to the staff at University of Maryland’s Amish Research Clinic and to the members of the Old Order Amish Community in Lancaster, PA who participated in this clinical trial.

## Data Availability

Primary data will be made available upon request to qualified academic investigators for research purposes under a Data Transfer Agreement to protect research participants’ confidential information.

